# Going beyond clinical routine in SARS-CoV-2 antibody testing - A multiplex corona virus antibody test for the evaluation of cross-reactivity to endemic coronavirus antigens

**DOI:** 10.1101/2020.07.17.20156000

**Authors:** Matthias Becker, Monika Strengert, Daniel Junker, Tobias Kerrinnes, Philipp D. Kaiser, Bjoern Traenkle, Heiko Dinter, Julia Häring, Anne Zeck, Frank Weise, Andreas Peter, Sebastian Hörber, Simon Fink, Felix Ruoff, Tamam Bakchoul, Armin Baillot, Stefan Lohse, Markus Cornberg, Thomas Illig, Jens Gottlieb, Sigrun Smola, André Karch, Klaus Berger, Hans-Georg Rammensee, Katja Schenke-Layland, Annika Nelde, Melanie Märklin, Jonas S. Heitmann, Juliane S. Walz, Markus Templin, Thomas O. Joos, Ulrich Rothbauer, Gérard Krause, Nicole Schneiderhan-Marra

**Author notes:** These authors contributed equally to this work (Matthias Becker, Monika Strengert, Ulrich Rothbauer). Corresponding author: Dr. Nicole Schneiderhan-Marra Markwiesenstrasse 55, 72770 Reutlingen, Germany.

## Abstract

Given the importance of the humoral immune response to SARS-CoV-2 as a global benchmark for immunity, a detailed analysis is needed to monitor seroconversion in the general population, understand manifestation and progression of COVID-19 disease, and ultimately predict the outcome of vaccine development. In contrast to currently available serological assays, which are only able to resolve the SARS-CoV-2 antibody response on an individual antigen level, we developed a multiplex immunoassay, for which we included spike and nucleocapsid proteins of SARS-CoV-2 and the endemic human coronaviruses (NL63, OC43, 229E, HKU1) in an expanded antigen panel. Compared to three commercial *in vitro* diagnostic tests, our MULTICOV-AB assay achieved the highest sensitivity and specificity when analyzing a well-characterized sample set of SARS-CoV-2 infected and uninfected individuals. Simultaneously, high IgG responses against endemic coronaviruses became apparent throughout all samples, but no consistent cross-reactive IgG response patterns could be defined. In summary, we have established and validated, a robust, high-content-enabled, and antigen-saving multiplex assay MULTICOV-AB, which is highly suited to monitor vaccination studies and will facilitate epidemiologic screenings for the humoral immunity toward pandemic as well as endemic coronaviruses.

## Introduction

Given the importance of the humoral immune response to SARS-CoV-2 as a global benchmark for immunity, a detailed analysis is needed to (i) monitor seroconversion in the general population^1,2^, (ii) understand manifestation and progression of the disease^3,4^, and (iii) predict the outcome of vaccine development^5,6^. Currently available serological assays utilize single analyte technologies such as ELISA to measure antibodies against SARS-CoV-2 antigens including spike (S) or nucleocapsid (N) protein^1,6-8^. To measure individual antibody (IgG and IgA) responses against SARS-CoV-2 and the endemic human coronaviruses (hCoVs) NL63, 229E, OC43, and HKU1, we developed a multiplexed immunoassay (MultiCoV-Ab), for which we included S and N proteins of these coronaviruses in an expanded antigen panel. Compared to commercial *in vitro* diagnostic (IVD) tests our MultiCoV-Ab assay achieved the highest sensitivity and specificity when analyzing 310 SARS-CoV-2 infected and 866 uninfected individuals. Simultaneously we see high IgG responses against hCoVs throughout all samples, whereas no consistent cross reactive IgG response patterns can be defined. In summary, our MultiCoV-Ab assay is highly suited to monitor vaccination studies and will facilitate epidemiologic screenings for the humoral immunity toward pandemic as well as endemic coronaviruses.

## Results

To investigate the antibody response of SARS-CoV-2-infected individuals, we developed and established a high-throughput and automatable bead-based multiplex assay, termed MultiCoV-Ab. We expressed and immobilized six different SARS-CoV-2-specific antigens on Luminex MAGPLEX beads with distinct color codes, specifically the trimeric full-length Spike protein (Spike Trimer), receptor binding domain (RBD), S1 domain (S1), S2 domain (S2), full-length nucleocapsid (N) and the N-terminal domain of nucleocapsid (N-NTD) (**Extended Data Fig. 1**). Immunoglobulins from serum and plasma samples were detected using phycoerythrin-labelled anti-human IgG or IgA antibodies. Data on quality control and assay performance is provided in **Extended Data Fig. 2** and **Extended Data Table 1**.

As key antigens for the classification of SARS-CoV-2-induced seroconversion, we used Spike Trimer and RBD previously described by Amanat *et. al*^2^, and initially screened a set of 205 SARS-CoV-2-infected and 72 uninfected individuals with the MultiCoV-Ab assay. Using a combined cut-off of both antigens, we identified all uninfected samples as negative (**Fig. 1a**). Of the 205 infected samples, the MultiCoV-Ab assay identified 24 (11.7%) as IgG antibody-negative. This finding is supported by three other commercially available IVD tests (Roche^9^, Siemens Healthineers^10^, Euroimmun^11^) widely used in clinical routine SARS-CoV-2 antibody testing. However, the IVD tests missed additional 8 (Roche), 11 (Siemens Healthineers) and 9 (Euroimmun) samples of SARS-CoV-2-infected individuals. Furthermore, the Euroimmun test classified 8 additional samples as “borderline” (**Fig. 1b, Extended Fig. 3a-c**). In accordance with our MultiCoV-Ab assay, no samples were classified as false positives by the Roche and Siemens tests, whereas one sample was classified as false positive and one as “borderline” by the Euroimmun test.

**Fig. 1.**
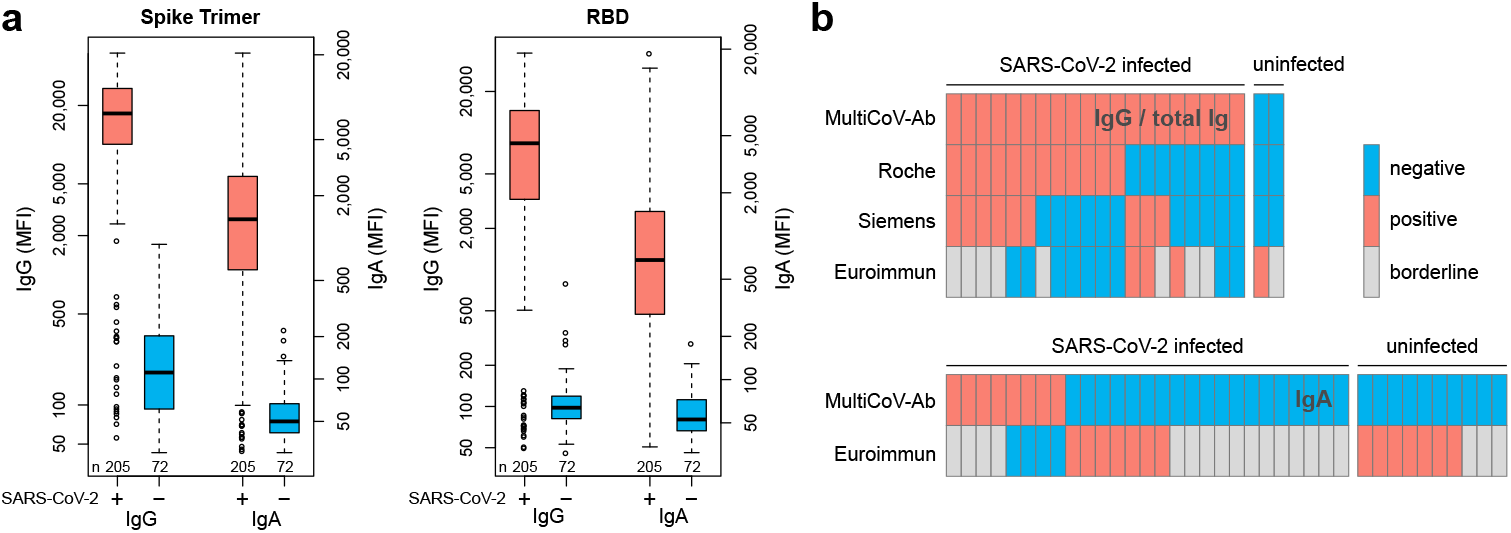
MultiCoV-Ab assay, a sensitive and specific tool to monitor SARS-CoV-2 antibody responses. **a**, Control sera (blue, n = 72) and sera from individuals with PCR-confirmed SARS-CoV-2 infection (red, n = 205) were screened in a multiplex bead-based assay using Luminex technology (MultiCoV-Ab) to quantify IgG or IgA responses to various antigens. Reactivity towards trimeric SARS-CoV-2 spike protein (Spike Trimer) or SARS-CoV-2 receptor binding domain of spike (RBD) was found to be the best predictor of SARS-CoV-2 infection. Data are presented as Box-Whisker plots of sample median fluorescence intensity (MFI) on a logarithmic scale. Outliers determined by 1.5 times IQR of log-transformed data are depicted as circles. **b**, Sample set from **a**, was used to compare assay performance of the MultiCoV-Ab assay using combined Spike Trimer and RBD antigens with commercially available single analyte SARS-CoV-2 assays which detect total Ig (Elecsys Anti-SARS-CoV-2 (Roche); ADVIA Centaur SARS-CoV-2 Total (COV2T) (Siemens Healthineers)) or IgG (Anti-SARS-CoV-2-ELISA - IgG (Euroimmun)) or IgA (Anti-SARS-CoV-2-ELISA - IgA (Euroimmun)). SARS-CoV-2 infection status of samples is indicated as SARS-CoV-2 positive (PCR) or negative. Antibody test results were classified as negative (blue), positive (red) or borderline (grey) as per the manufacturer’s definition. Only samples with divergent antibody test results are shown.

When testing for IgA antibodies in samples of SARS-CoV-2-infected individuals, our MultiCoV-Ab assay classified 47 (22.9 %) as IgA-negative, whereas the Euroimmun test classified 32 (15.6 %) as IgA-negative, and 16 (7.8 %) as borderline (**Fig. 1b and Extended Data Fig. 3d**). For the uninfected samples, the Euroimmun test identified 7 (9.7 %) as false positives and 3 (4.2 %) as “borderline”, whereas no samples were classified as false positives by the MultiCoV-Ab assay.

Next, we used an extended sample set with 310 SARS-CoV-2-infected and 866 uninfected donors for clinical validation of MultiCoV-Ab assay. A simplified overview of this set is shown in **Fig. 2a**; a complete breakdown is displayed in **Extended Data Table 2**. A direct comparison revealed that Spike Trimer and RBD were the best predictors of SARS-CoV-2 infection. Signal cut-offs were defined for both, IgG and IgA detection, based on ROC analysis with focus on maximum specificity. Additionally, we defined a cut-off for overall IgG and IgA positivity for which both individual cut-offs for Spike Trimer and RBD had to be met (**Fig. 2b**). As shown above, cut-offs based on IgG were shown to be more sensitive and specific than those based on IgA. With the IgG overall cut-off, we reached a specificity of 100 % (**Fig. 2c**), which would not have been possible for either of the antigens individually, while still retaining acceptable sensitivity. To identify samples with an early immune response, we simultaneously measured IgA response. With the MultiCoV-Ab assay, we identified eight IgA-positive samples that showed no IgG response (**Fig. 2d**). Two of these were uninfected and falsely classified as positive. For four of the remaining six infected samples, details regarding the time between the onset of symptoms and sample drawing were available (2, 6, 7, and 15 days). We hypothesized that IgA in these samples can be used to measure an early onset of antibody response. Thus, we classified samples with strong IgA positivity - signal to cut-off (S/CO) > 2 for Spike Trimer and RBD - as “positive”, irrespective of their detected IgG response. With this combined IgG + IgA classification, we reached an optimal sensitivity of 90 % while retaining a specificity of 100 %.

**Fig. 2.**
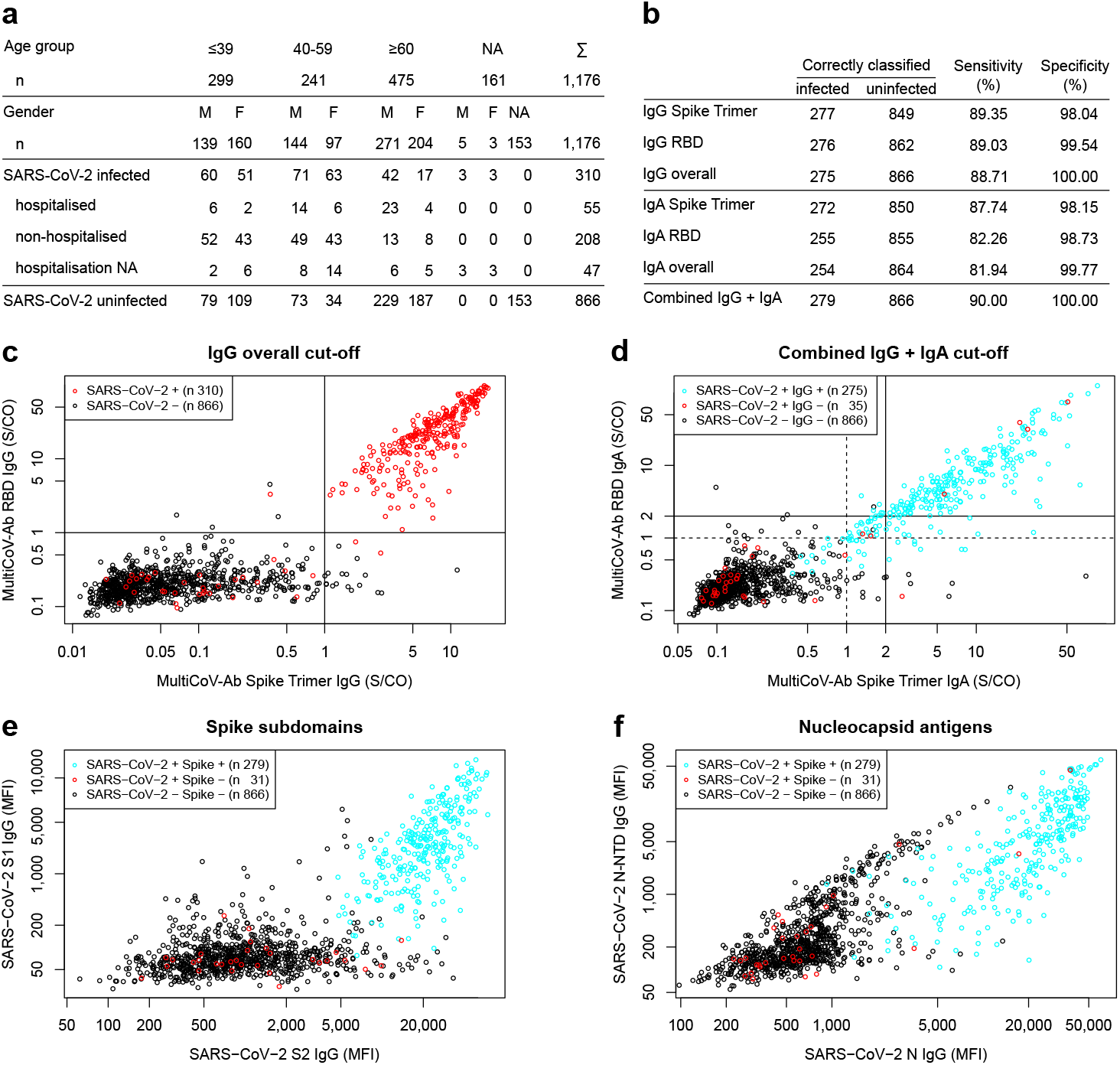
Combination of 2 Spike protein variants and isotype profiling by multiplex assay increases accuracy to identify SARS-CoV-2 antibody positive individuals. **a**, An extended sample set of SARS-CoV-2-uninfected (n = 866) and SARS-CoV-2-infected individuals (n = 310) was used to further validate our MultiCoV-Ab assay. Age, gender, SARS-CoV-2 infection status and hospitalization status of study population are shown. NA: information was not provided. **b**, MultiCoV-Ab assay sensitivity and specificity were calculated for IgA or IgG based on a single analyte or a combined cut-off of Spike Trimer and RBD (IgG or IgA overall). A cut-off combining IgG and IgA was calculated as well. **c-d**, Scatterplot detailing MultiCoV-Ab assay cut-offs. Signal to cut-off (S/CO) values are displayed for Spike Trimer against RBD on a logarithmic scale. For IgG (**c**,), cut-offs are visualized by straight lines and SARS-CoV-2-infected and uninfected samples are separated by color (black circles – SARS-CoV-2-uninfected; red circles – SARS-CoV-2-infected). For IgA (**d**,) cut-offs are visualized as dashed lines and S/CO of 2 used for the combined cut-off is shown as straight lines. SARS-CoV-2-infected samples are split into IgG-positives and - negatives by color as indicated in the plot. **e-f**, Scatterplots display IgG response to additional SARS-CoV-2 antigens contained in the MultiCoV-Ab panel: MFI for Spike subdomains S1 vs S2 (**e**,) or Nucleocapsid antigens N vs N-NTD (**f**,) are displayed on logarithmic scale. SARS-CoV-2-uninfected samples are distinguished from SARS-CoV-2-infected and MultiCoV-Ab classification into positives or negatives as indicated by color.

Further analyzing the Ig response towards both subdomains of the spike, S1 and S2, we achieved no additional sensitivity for the classifier (**Fig. 2e**). Interestingly, RBD, as a part of S1, showed much fewer uninfected samples with increased IgG response compared to S1. For S2 even more uninfected samples had increased signals, pointing to potential cross-reactivity in this domain of the spike protein (**Fig. 2e**). We further complemented our assay with the N and N-NTD proteins. Although these antigens were successfully used in single-analyte assays^12^, we observed a high cross reactivity in uninfected samples for both (**Fig. 2f**). Across the entire data set, only one sample showed a distinct immune response to N and N-NTD, but not to all spike derived antigens.

Longitudinal samples from five hospitalized patients were subjected to a small-scale time course analysis of IgG and IgA immune responses (**Fig. 3a**). Levels of both Ig classes strongly increased within the first ten days after the onset of symptoms. While IgG levels appeared constant over roughly two months, IgA levels started to decline between day 10 and 20 after the onset of symptoms where samples were available. These effects were consistent for the majority of SARS-CoV-2 antigens. Furthermore, we found that patients’ hospitalization, as a measure of disease severity (**Fig. 3b**), seemed to correlate with an increased humoral immune response, especially in IgA. Furthermore, there is indication for a trend for increasing age as well (**Fig. 3c**). However, it should be considered that patients of higher age also had a higher rate of hospitalization in our study population.

**Fig. 3.**
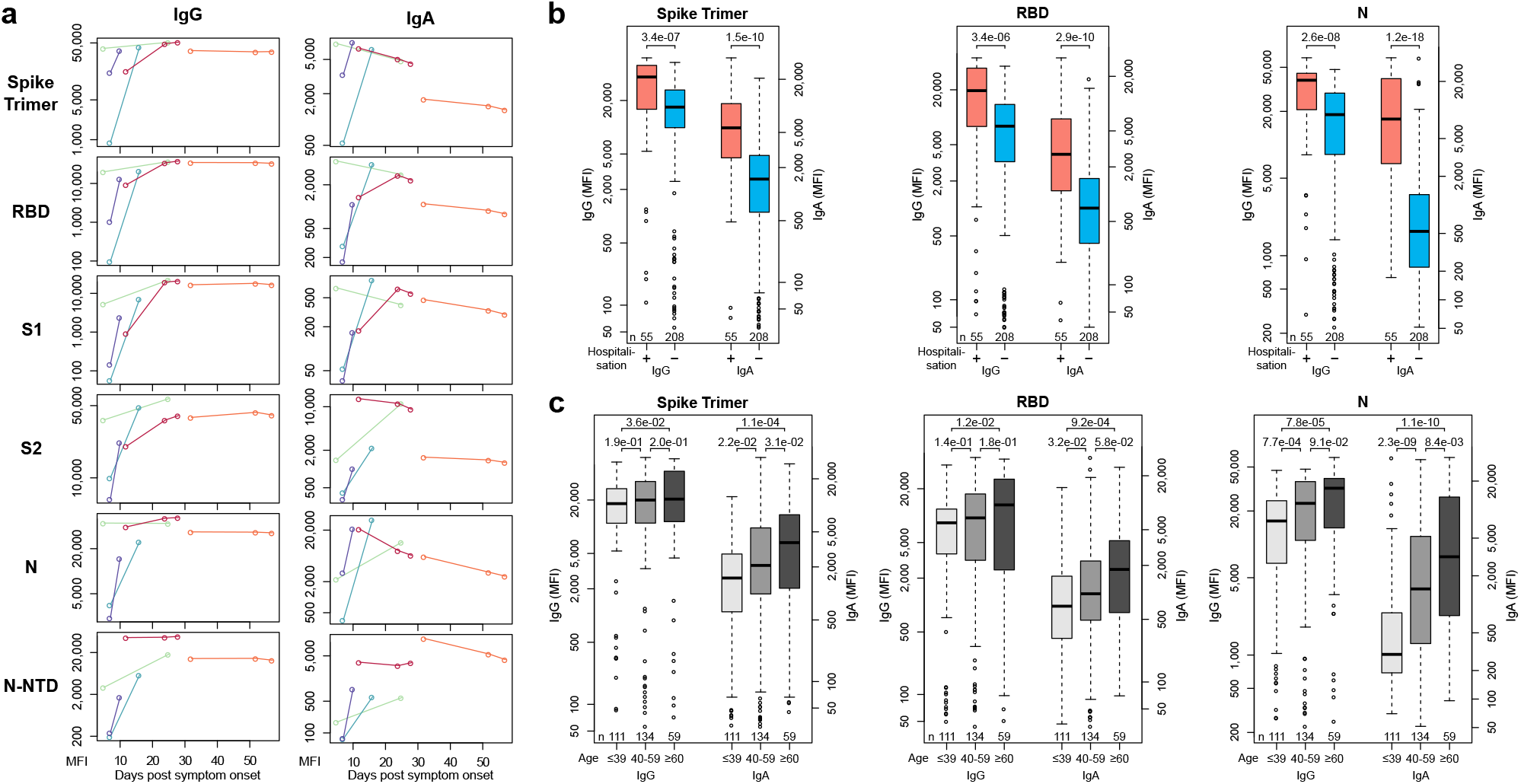
Multiplex-based seroprofiling allows in-depth characterization of SARS-CoV-2 antibody responses. **a**, Kinetic of SARS-CoV-2 antigen-specific IgA and IgG responses is shown for indicated days after symptom onset for six SARS-CoV-2-specific antigens for five different patients. **b-c**, Samples of SARS-CoV-2-infected individuals were analysed to identify antigen-and isotype-specific antibody responses based on hospitalization indicating disease severity (**b**,) or age (**c**,). Data is presented as Box-Whisker plots of sample MFI on a logarithmic scale. Outliers determined by 1.5 times IQR of log-transformed data are depicted as circles. p-value (Mann-Whitney U test, two-sided) is displayed at the top of the boxes, indicating differences between signal distribution for respective groups.

In order to explore cross-reactivity of hCoVs with SARS-CoV-2, we included S1, N, and N-NTD antigens from human α-(NL63 and 229E) and β-hCoVs (OC43 and HKU1) in our MultiCoV-Ab panel (**Extended Data Fig. 1**). The immune response towards all hCoV antigens was more dependent on coronavirus clade than on N or S1 antigen. However, within the clades of α-hCoVs and β-hCoVs, types of antigens were more dominant than the virus subtype, as demonstrated by rank correlation analysis and hierarchical clustering. Interestingly, IgG response against α-hCoVs clustered more closely to SARS-CoV-2 than to β-hCoVs (**Fig. 4a, Extended Data Fig. 4a**). Overall, we identified a considerable immune response to hCoV antigens throughout the whole sample set with no notable differences between samples from SARS-CoV-2-infected and uninfected donors in IgG or IgA for S1 (**Fig. 4b**), N (**Fig. 4c**), or N-NTD (**Extended Data Fig. 4b**).

**Fig. 4.**
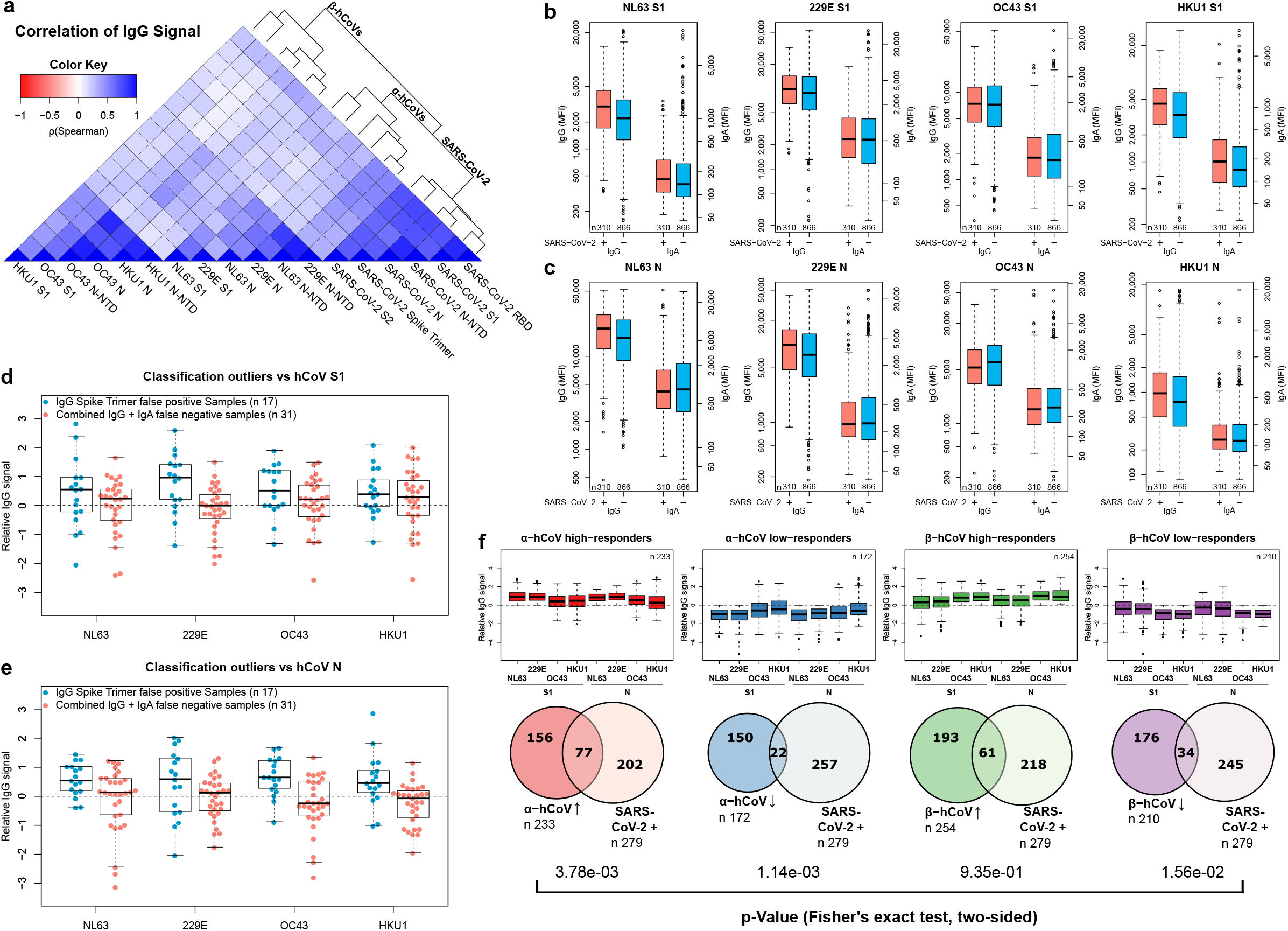
Correlation of endemic and SARS CoV-2 antibody responses. **a**, Correlation of IgG response for the entire sample set (n = 1176) is visualized as heatmap based on Spearman’s ρ coefficient; dendrogram on the right side displays antigens after hierarchical clustering was performed. **b-c**, Immune responses (IgG and IgA) towards hCoV S1 (**b**,) and N (**c**,) proteins are presented as Box-Whisker plots of sample MFI on a logarithmic scale for SARS-CoV-2-infected (red, n = 310) and uninfected (blue, n = 866) individuals. Outliers determined by 1.5 times IQR of log-transformed data are depicted as circles. **d-e**, Relative levels of IgG-specific immune response towards hCoV S1 (**d**,) and N (**e**,) proteins are presented as Box-Whisker plots / stripchart overlays of log-transformed and per-antigen scaled and centred MFI for the sample subsets of Spike Trimer false positives (blue, n =17) and combined IgG + IgA false negatives (red, n = 31). **f**, From the entire study population, groups of α-or β-hCoV high and low responders were built as indicated. High responder were defined as samples with above average MFI values for S1 and N-specific IgGs of the respective hCoV clade. Low responders were defined with below MFI values, correspondingly. Responder groups (i) α-hCoV ↑, red, n = 233, (ii) β-hCoV ↑, green, n = 254, (iii) α-hCoV ↓, blue, n = 172 (iv) β-hCoV ↓, purple, n = 210 are shown as Box-Whisker plots of log-transformed and per-antigen scaled and centred MFI values across hCoV N and S1 antigens. Outliers determined by 1.5 times IQR are depicted as circles. The over-or under-representation of SARS-CoV-2 responders (SARS-CoV-2 +, n = 279, as determined by positive MultiCoV-Ab classification) within the four sample groups is visualized in Venn diagrams, stochastic significance was calculated using Fisher’s exact test (two-sided).

We therefore used the IgG signal relative to the average response per antigen for further analyses, which allowed comparison among all hCoV antigens on one scale. For those uninfected samples, which showed an IgG cross reactivity towards Spike Trimer (Spike Trimer false positives), we observed partially increased responses towards hCoV antigens. Those samples, which did not show an immune response after SARS-CoV-2 infection (false negatives, as determined by MultiCoV-Ab assay, combined IgG + IgA) were closer to the baseline (**Fig. 4d-e, Extended Data Fig. 4c**). This indicates that cross-reactivity with hCoVs causes some of the observed SARS-CoV-2 immune response in samples taken from individuals not exposed to SARS-CoV-2.

To investigate the correlation of hCoV and SARS-CoV-2 immune response further, we grouped samples into high and low responders for α-hCoVs and β-hCoVs. High responders had relative IgG signals > 0 for N and S1 antigens of both hCoV subtypes within the clade, while low-responders had < 0, respectively. Samples with SARS-CoV-2 immune response (as determined by MultiCoV-Ab assay, combined IgG + IgA classification) were significantly overrepresented within the group of α-hCoV high responders (p = 3.78e-03, Fisher’s exact test, two sided), while being significantly underrepresented within the group of α-hCoV and β-hCoV low responders (p = 1.14e-03 and p = 1.56e-02, respectively, Fisher’s exact test, two sided) (**Fig. 4f**). These results showed that while there were no discernible global effects for single antigens, there is a correlation between the SARS-CoV-2 immune response with high hCoV responses, especially towards α-hCoVs.

## Discussion

We demonstrated that our MultiCoV-Ab, a novel multiplex assay, is highly suitable to classify seroconversion in SARS-CoV-2-infected individuals. With a combined cut-off using SARS-CoV-2 trimeric full-length spike protein and RBD, we were able to eliminate false positive responses and achieved a sensitivity of 90% with a specificity of 100% for 310 samples from SARS-CoV-2-infected and for 866 samples from uninfected individuals. We found that detection of IgG more accurately reflected infection compared to IgA, although both were highly specific. However, by simultaneously monitoring IgA, we additionally were able to detect an early immune response in some patients. The MultiCoV-Ab approach allows the easy addition of SARS-CoV-2-specific antigens, here six in total, which provides an additional level of confidence in patient classification. Thus, for example, we noticed that the spike S1 domain showed fewer false positive responses compared to the S2 domain. Interestingly, Ng *et al*.^13^ reported reactivity towards SARS-CoV-2 S2 from sera of patients with recent seasonal hCoV infection. These sera prevented infection with SARS-CoV-2 pseudotypes in a neutralization assay. Additionally, we found that spike non-responders also did not show a response to nucleocapsid, which has been described as strongest inducer of antibody responses^12,14^; and not vice versa.

In our comparison to commercially available IVD tests, we classified fewer samples as false negative using our MultiCoV-Ab assay. For 10% of all infected samples, we could not detect a SARS-CoV-2 specific immune response, which is in line with previous findings^3,15,16^. Those non-responders may be able to limit viral replication by innate immune mechanisms or cellular immunity is dominant in mediating viral clearance^17,18^.

Expanding our MultiCoV-Ab assay to the endemic hCoVs NL63, 229E, OC43, and HKU1 revealed a clear IgG immune response for all tested samples. Furthermore, we did not observe a difference for the samples from proven hCoV-infected individuals, compared to other samples. Due to the general lack of availability of samples from hCoV-naïve individuals, it was difficult to analyze hCoV-mediated cross-reactivity. Nevertheless, our multiplexed readout indicates a correlation between the SARS-CoV-2 immune response and high hCoV responses. Currently, we are identifying population groups which were highly exposed and showed different susceptibility to SARS-CoV-2 infection, e.g. the “Ischgl-study group” (unpublished data)^19^, in order to elucidate potential cross protection derived from immune responses towards endemic hCoVs in more detail. Alternatively, studies analyzing hCoV signatures in samples from individuals before and after SARS-CoV-2 infection using the MultiCoV-Ab assay would help to get insight into a potential cross protection.

A multiplex setup such as MultiCoV-Ab assay is especially suited to vaccination studies, since the flexibility and broad antigen coverage allows to efficiently map vaccine immune responses to an immunoglobulin isotype and subtype level for the target pathogen and related species ^20^. Interestingly, previous SARS-CoV-1 vaccine studies clearly indicated that a detailed characterization of vaccine-induced antibody responses is mandatory for efficient coronavirus vaccine development^21,22^.

In summary, we have established and validated the MultiCoV-Ab assay, a robust, high-content-enabled, and antigen-saving multiplex assay. This assay is suitable for comprehensive characterization of SARS-CoV-2 infection on the humoral immune response and for epidemiological screenings to accurately measure SARS-CoV-2 seroprevalence in large cohort studies. It further provides the unique opportunity to assess and correlate immunity for both endemic and pathogenic coronaviruses. Finally, the multiplex nature of the MultiCoV-Ab assay can deliver urgently needed data on the outcome of SARS-CoV-2 vaccination.

## Data Availability

Raw data and analysis files are available for download as supplemental files of this manuscript

## Supplementary - Materials and Methods

### Generation of expression constructs for production of viral antigens

The cDNAs encoding the full-length nucleocapsid proteins of SARS-CoV-2, hCoV-OC43, hCoV-NL63, hCoV-229E, and hCoV-HKU1 (GenBank accession numbers QHD43423.2; YP_009555245.1; YP_003771.1; NP_073556.1; YP_173242.1) were produced with an N-terminal hexahistidine (His_6_)-tag by DNA synthesis (ThermoFisher Scientific). The cDNAs were cloned by standard techniques into NdeI/HindIII sites of the bacterial expression vector pRSET2b (ThermoFisher Scientific). To generate N-terminal domains (NTDs) of the respective nucleocapsid proteins (SARS-CoV-2 NTD aa 1-189; hCoV-OC43 NTD aa 1-204; hCoV-NL63 NTD aa 1 - 154; hCoV-229E NTD aa 1-156; hCoV-HKU1 NTD aa 1-203), a stop codon located N-terminally to the Serine-Arginine (SR)-rich linker site^23^ was introduced via PCR mutagenesis of the nucleocapsid encoding plasmids using the forward primer pRSET2b down-for 5’ - GGT AAG CTT GAT CCG GCT GCT AA - 3’ and respective reverse primers: SARS-CoV2_NTD-rev 5’ - GGG AAG CTT ACT CAG CAT AGA AGC CCT TTG G - 3’, OC43_NTD-rev 5’ - GGG AAG CTT ATT CGA TAT AAT AGC CCT GCG G - 3’, NL63_NTD-rev 5’ - GGG AAG CTT ATT CAA CAA CGC TCA GTT CCG - 3’, 229E_NTD-rev 5’ - GGG AAG CTT ATT CAA CAA CGG TAA CAC CAT TC - 3’ and HKU1_NTD-rev 5’ - GGG AAG CTT ATT CCA CAT AGT AGC CCT GAG GC-3’.

The pCAGGS plasmids encoding the stabilized trimeric Spike protein and the receptor binding domain (RBD) of SARS-CoV-2 were kindly provided by F. Krammer^2^.

The cDNA encoding the S1 domain (aa 1 - 681) of the SARS-CoV-2 Spike protein was obtained by PCR amplification using the forward primer S1_CoV2-for 5’-CTT CTG GCG TGT GAC CGG - 3’ and reverse primer S1_CoV2-rev 5’ - GTT GCG GCC GCT TAG TGG TGG TGG TGG TGG TGG GGG CTG TTT GTC TGT GTC TG - 3’ and the full length SARS-CoV-2 SPIKE cDNA as template and cloned into the XbaI/NotI-digested backbone of the pCAGGS vector, thereby adding a C-terminal His_6_-Tag.

The cDNAs encoding the S1 domains of hCoV-OC43 (aa 1 - 760), hCoV-NL63 (aa 1 - 744), hCoV-229E (aa 1 - 561) and hCoV-HKU1 (aa 1 - 755) (GenBank accession numbers AVR40344.1; APF29071.1; YP_003771.1; APT69883.1; AGW27881.1) were produced by DNA synthesis (ThermoFisher Scientific), digested using XbaI/NotI and ligated into the pCAGGS vector. All expression constructs were verified by sequence analysis.

### Protein expression and purification

For expression of the viral nucleocapsid proteins (full-length nucleocapsid and N-NTDs), the respective expression constructs were used to transform *E.coli* BL21 (DE3) cells. Protein expression was induced in 1 L TB medium at an optical density (OD_600_) of 2.5 - 3 by addition of 0.2 mM isopropyl-β-D-thiogalactopyranoside (IPTG) for 16 h at 20 °C. Cells were harvested by centrifugation (10 min at 6,000 x g) and the pellets were suspended in binding buffer (1x PBS, ad 0.5 M NaCl, 50 mM imidazole, 2 mM phenylmethylsulfonyl fluoride, 2 mM MgCl_2_, 150 µg/mL lysozyme (Merck) and 625 µg/mL DNaseI (Applichem)). Cell suspensions were sonified for 15 min (Bandelin Sonopuls HD70 - power MS72/D, cycle 50%) on ice, incubated for 1 h at 4 °C in a rotary shaker followed by a second sonification step for 15 min. After centrifugation (30 min at 20,000 x g), urea was added to a final concentration of 6 M to the soluble protein extract. The extract was filtered through a 0.45 µm filter and loaded on a pre-equilibrated 1-mL HisTrap^FF^ column (GE Healthcare). The bound His-tagged nucleocapsid proteins were eluted by a linear gradient (30 mL) ranging from 50 to 500 mM imidazole in elution buffer (1x PBS, pH 7.4, 0.5 M NaCl, 6 M urea). Elution fractions (0.5 mL) containing the His-tagged nucleocapsid proteins were pooled and dialyzed (D-Tube Dialyzer Mega, Novagen) against PBS.

The viral S1-domains, SARS-CoV-2 RBD, and the stabilized trimeric SARS-CoV-2 Spike protein were expressed in Expi293 cells following the protocol as described in Stadlbauer *et al*.^8^.

All purified proteins were analyzed via standard SDS-PAGE followed by staining with InstantBlue Coomassie stain (Expedeon) and immunoblotting using an anti-His antibody (Penta-His Antibody, #34660, Qiagen) in combination with a donkey-anti-mouse antibody labeled with AlexaFluor647 (Invitrogen) on a Typhoon Trio (GE-Healthcare, Freiburg, Germany; excitation 633 nm, emission filter settings 670 nm BP 30) to confirm protein integrity. To further confirm correct expression, integrity, and purity, proteins were analysed by mass spectrometry. To control the production reproducibility of the antigens, potential aggregation and melting temperatures of the proteins were investigated by nano differential scanning fluorimetry (nanoDSF) using a Prometheus (Nanotemper, Munich, Germany).

### Commercial antigens

Two commercial antigens were used to complement the in-house-produced antigen panel. The S2 ectodomain of the SARS-CoV-2 spike protein (aa 686 – 1213) was purchased from Sino Biological, Eschborn, Germany (cat # 40590, lot # LC14MC3007). A full-length nucleocapsid protein of SARS-CoV-2 was purchased from Aalto Bioreagents, Dublin, Ireland (cat # 6404-b, lot # 4629).

### Bead-based serological multiplex assay

All antigens were covalently immobilized on spectrally distinct populations of carboxylated paramagnetic beads (MagPlex Microspheres, Luminex Corporation, Austin, TX) using 1-ethyl-3-(3-dimethylaminopropyl)carbodiimide (EDC)/ sulfo-N-hydroxysuccinimide (sNHS) chemistry. For immobilization, a magnetic particle processor (KingFisher 96, Thermo Scientific, Schwerte, Germany) was used.

Bead stocks were vortexed thoroughly and sonicated for 15 seconds. Subsequently, 83 µL of 0.065% (v/v) Triton X-100 and 1 mL of bead stock containing 12.5 x 10^7^ beads of one single bead population were pipetted into each well. The beads were then washed twice with 500 µL of activation buffer (100 mM Na2HPO4, pH 6.2, 0.005% (v/v) Triton X-100) and beads were activated for 20 min in 300 µL of activation mix containing 5 mg/mL EDC and 5 mg/mL sNHS in activation buffer. Following activation, the beads were washed twice with 500 µL of coupling buffer (500 mM MES, pH 5.0, 0.005% (v/v) Triton X-100) and the antigens were added to the activated beads and incubated for 2 h at 21 °C to immobilize the antigens on the surface.

Antigen-coupled beads were washed twice with 800 µL of wash buffer (1x PBS, 0.005 % (v/v) Triton X-100) and were finally resuspended in 1,000 µL of storage buffer (1x PBS, 1 % (w/v) BSA, 0.05% (v/v) ProClin). The beads were stored at 4°C until further use.

To detect human IgG and IgA responses against SARS-CoV-2 and the endemic human coronaviruses (hCoV-NL63, hCoV-229E, hCoV-OC43 and hCoV-HKU1), the purified trimeric Spike protein (S), S1-domain, S2-domain (Sino Biological GmbH, Europe), RBD, nucleocapsid (N) and the N-terminal domain of nucleocapsid (N-NTD) of SARS-CoV-2 as well as the S1-domain, N, and N-NTD of the endemic hCoVs were immobilized on different bead populations as described above. The individual bead populations were combined into a bead mix. A bead-based multiplex assay was performed. Briefly, samples were incubated at a 1:400 dilution for 2 hours at 21 °C. Unbound antibodies were removed and the beads were washed three times with 100 µL of wash buffer (1x PBS, 0.05% (v/v) Tween20) per well using a microplate washer (Biotek 405TS, Biotek Instruments GmbH). Bound antibodies were detected with R-phycoerythrin labeled goat-anti-human IgG or IgA antibodies (incubation for 45 min at 21°C). Measurements were performed using a Luminex FLEXMAP 3D instrument and the Luminex xPONENT Software 4.3 (settings: sample size: 80 µL, 50 events, Gate: 7,500 – 15,000, Reporter Gain: Standard PMT).

### Data analysis

Data analysis and visualization was performed with R Studio (Version 1.2.5001, using R version 3.6.1) using the Median Fluorescent Intensity (MFI). Statistical analysis was performed using R package “stats” from the base repository. Mann-Whitney U test was used to determine difference between signal distributions from different sample groups. Spearman’s ρ coefficient was calculated in order to correlate antigens by response from the entire sample set, followed by hierarchical clustering to group antigens. Fishers’ exact test was used to calculate significance of overlap between sample groups.

### Quality control

In order to test the repeatability of the MultiCoV-Ab assay three quality control samples (QCs) were processed in duplicate on each test plate (n = 17) during the sample screening and inter-assay variance was assessed for each antigen in the multiplex. For intra-assay variance, 24 replicates for each of the three QC samples were analyzed on one plate. Achieved results are presented in **Extended Data Table 1 and Extended Data Fig. 2**. A limit of detection (LOD) for each antigen was determined by processing a blank in 24 replicates and LOD was set as mean MFI + 3 standard deviations. Sample parallelism and comparability of paired serum and plasma samples was assessed over eight dilution steps ranging from 1:100 to 1:12,800 (**Extended Data Fig. 2**). A set of samples derived from 205 SARS-CoV-2-infected and 72 uninfected individuals was tested repeatedly with two different kit batches. The samples classification in both runs matched 100 %.

### Samples

A total of 1176 sera and plasma samples were used for the MultiCoV-Ab assay development. Ethical approval was granted from the Ethics Committee of Hannover Medical School (#9122_BO_K2020). Only de-identified samples were used for the MultiCoV-Ab assay development. All samples were pre-existing. Cohort age was 5-88 years; age was not known for 161 samples.

310 samples were from COVID-19 patients or convalescents. Samples were classified as SARS-CoV-2 infected, if a positive SARS-CoV-2 RT-PCR was reported and/ or if hospitalization/ quarantine for COVID-19 was indicated as part of the samples metadata. ΔT defined as time between PCR test or symptom onset and blood draw was 0-73 days (median= 38 d; n=258). ΔT was not provided for 52 samples. SARS-CoV-2 infected samples used in this study were collected after ethical review (9001_BO_K, Hannover Medical School; 179/2020/BO2, University Hospital Tübingen; 85/20, Ärztekammer des Saarlandes).

866 control samples were from non-SARS-CoV-2 infected individuals and were classified as non-infected as they were obtained prior to the emergence of SARS-CoV-2 in December 2019 or because they were taken from individuals who had not reported cold symptoms since the beginning of 2020.

The majority of non-SARS-COV-2 infected samples was randomly selected and consistent of prepandemic blood donors, commercially available (Central BioHub GmbH, Berlin, Germany and BBI Solutions, Crumlin, UK) or bio-banked specimens. 365 samples were from the Memory and Morbidity in Augsburg Elderly (MEMO) study (a subcohort of the MONICA S2 cohort (WHO 1988)) and were included based on available serological titers for HSV-1, HSV-2, HHV-6 and EBV^24^. 88 samples were obtained from transplanted patients with chronic respiratory conditions.

Collection of non-SARS-CoV-2 infected control samples had been approved by several ethic votes: 3232-2016 (Ethics Committee of Hannover Medical School); 62/20 (Ethics Committees of the Medical Faculty of the Saarland University at the Saarland Ärztekammer); WUM 17.02.1997 (Joint ethics committee of the University of Muenster and the Westphalian Chamber of Physicians),

Additional sample details can be found in Extended Data Table 2.

### Data availability

Primary Data including raw MFI and sample annotation will be made available upon request.

### Code availability

R Code for data analysis will be made available upon request.

## Acknowledgements

This work has received funding from the European Union’s Horizon 2020 research and innovation programme under grant agreement No 101003480 – CORESMA as well as from the German Federal Ministry for Research and Education/the Helmholtz Association. We thank Florian Krammer providing us with expression plasmids for the Spike Trimer and RBD. We thank Shannon Layland for critical proofreading of the manuscript.

## Author Contributions

M.B. designed and performed experiments and data analysis; M.S. designed experiments; D.J., J.H., S.F., F.R., planned and performed experiments; A.Z. performed mass spectrometry analysis; J.H. performed nanoDSF analyses; H.D., B.T., P.K., F.W., U.R. designed, cloned, expressed and purified the antigens; S.H. and A.P. performed sample analysis; T.B., A.B., S.L., S.S., M.C., T.I., H-G.R., A.N., J.W., M.T., T.O.J. arranged sample and data collection; M.T., T.O.J, G.K. supported the study planning; N.S-M. planned the assay development and validation and designed experiments; M.B., M.S., U.R. and N.S-M. wrote the manuscript. All authors reviewed the manuscript.

## Extended Data Figure Legends

**Extended Data Fig. 1.**
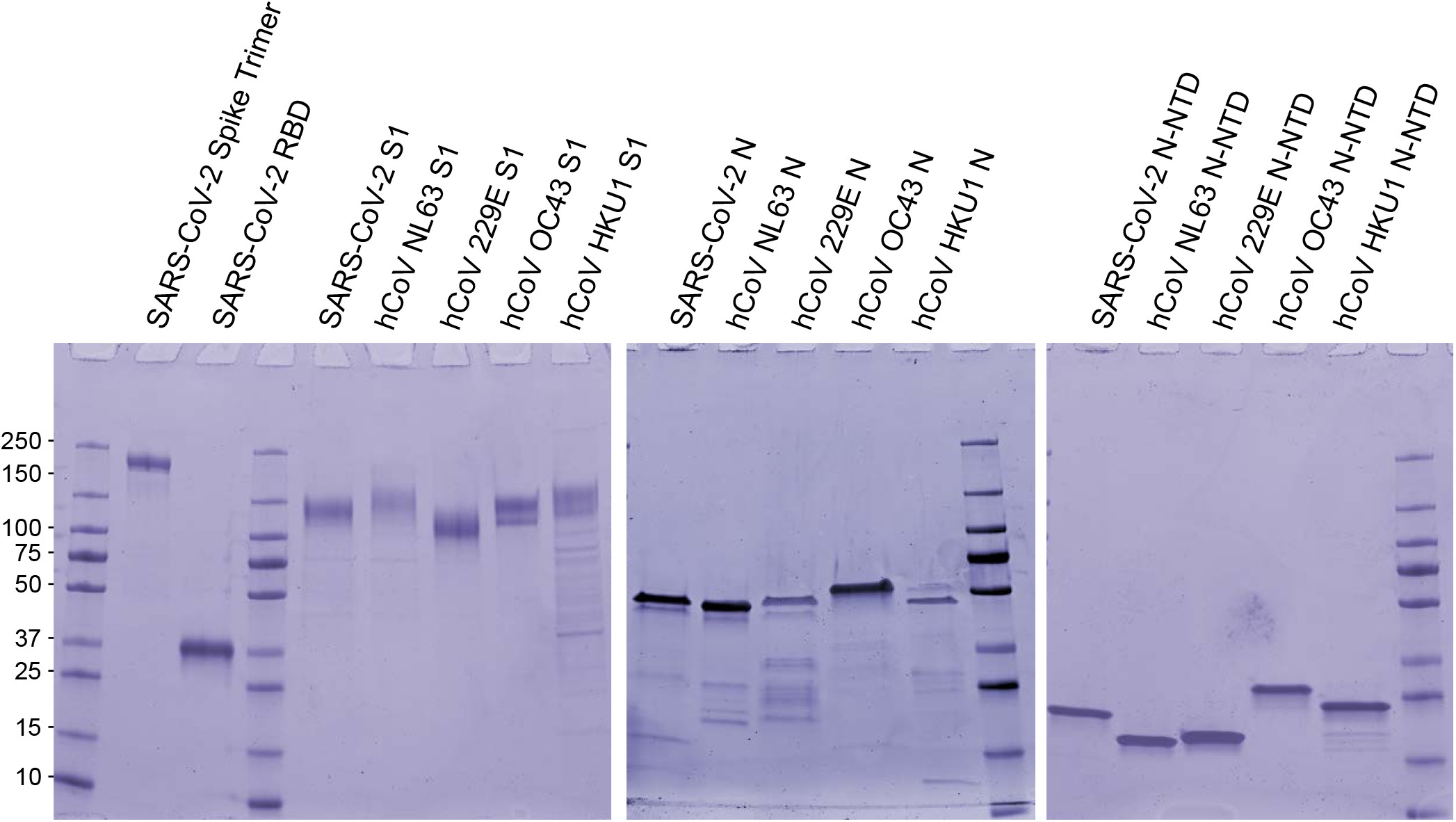
SDS-PAGE analysis of the recombinant viral antigens used in this study. To test for purity and integrity 1 - 2 µg of indicated recombinant proteins were boiled in reducing SDS-sample buffer and subjected to a gradient (4 - 20%) SDS-PAGE followed by coomassie staining. SARS-CoV-2_Spike, SARS-CoV-2_RBD and the S1-domains of SARS-CoV-2, hCoV-NL63, hCoV-229E, hCoV-OC43 and hCoV-HKU1 were produced in ExpiHEK™ cells. Nucleocapsid (N) and N-terminal domain of nucleocapsid (N-NTD) of SARS-CoV-2, hCoV-NL63, hCoV-229E, hCoV-OC43 and hCoV-HKU1 were produced in *E.coli*.

**Extended Data Fig. 2.**
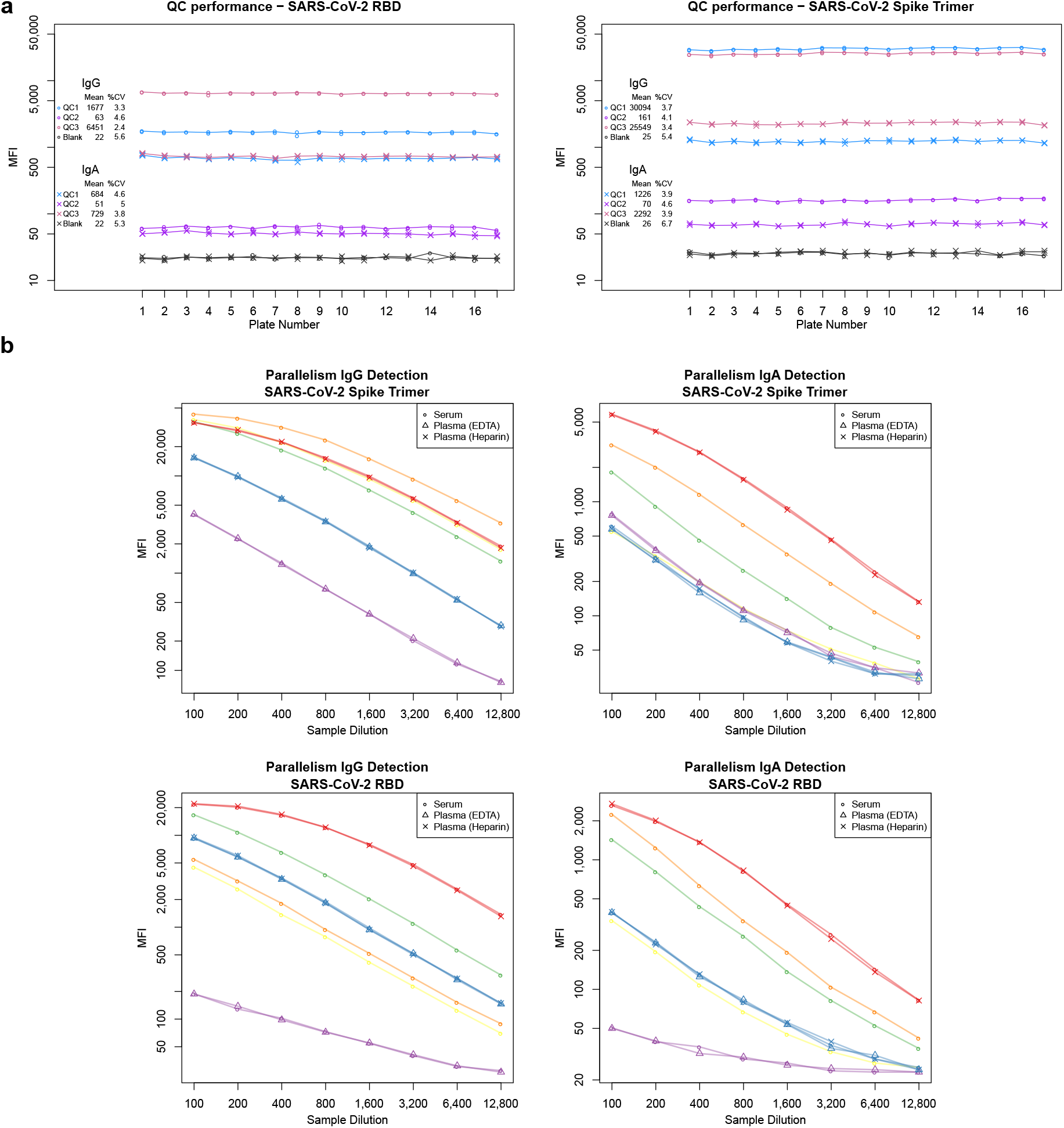
**a**, Three quality control (QC) samples, as well as a sample of assay buffer (blank sample) were processed in duplicates on every plate. Performance across 17 assay runs is depicted and mean and %CVs are shown on the left side. For plate 14, a processing error lead to exclusion of one blank sample from this evaluation. **b**, To assess parallelism of signals from different samples, 6 unique serum samples were processed over a dilution series of 8 steps from 1:100 to 1:12,800. For 3 samples, paired plasma (EDTA and/or Heparin) were available and processed together. For IgG and IgA detection of Spike Trimer and RBD, MFI are plotted against sample dilution. Color indicates unique sample and shapes indicate sample type.

**Extended Data Fig. 3.**
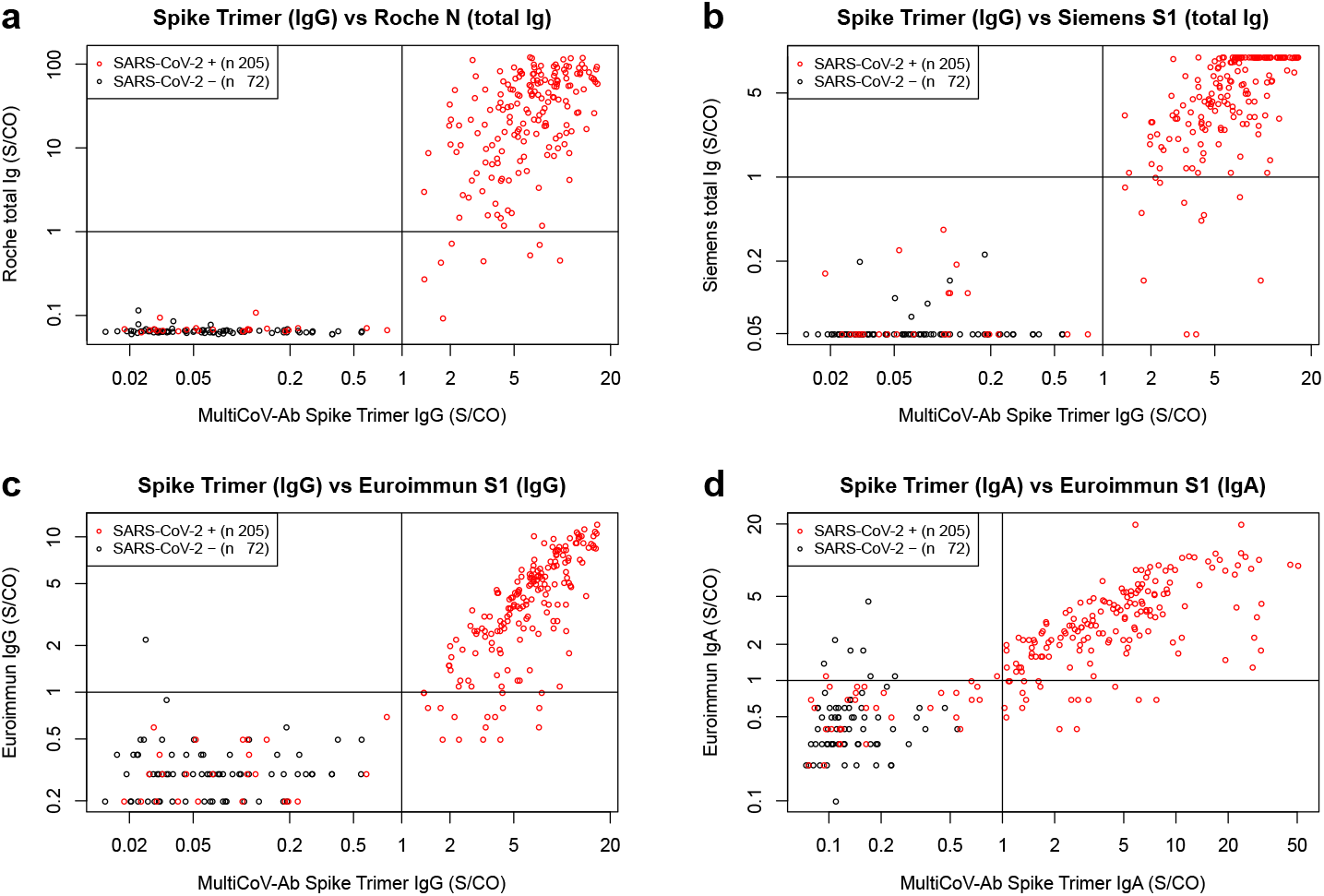
**a-d**, Scatterplots of sample set with defined SARS-CoV-2 infection status (infected: red, n=205; uninfected: black, n=72) to compare performance of the MultiCoV-Ab Spike Trimer vs indicated antigens of commercial SARS–CoV-2 test kits. Signals are depicted as Signal to cut-off ratios (S/CO) on a logarithmic scale. Lines indicate the respective cut-off values as defined by the manufacturer to determine positive and negative test results.

**Extended Data Fig. 4.**
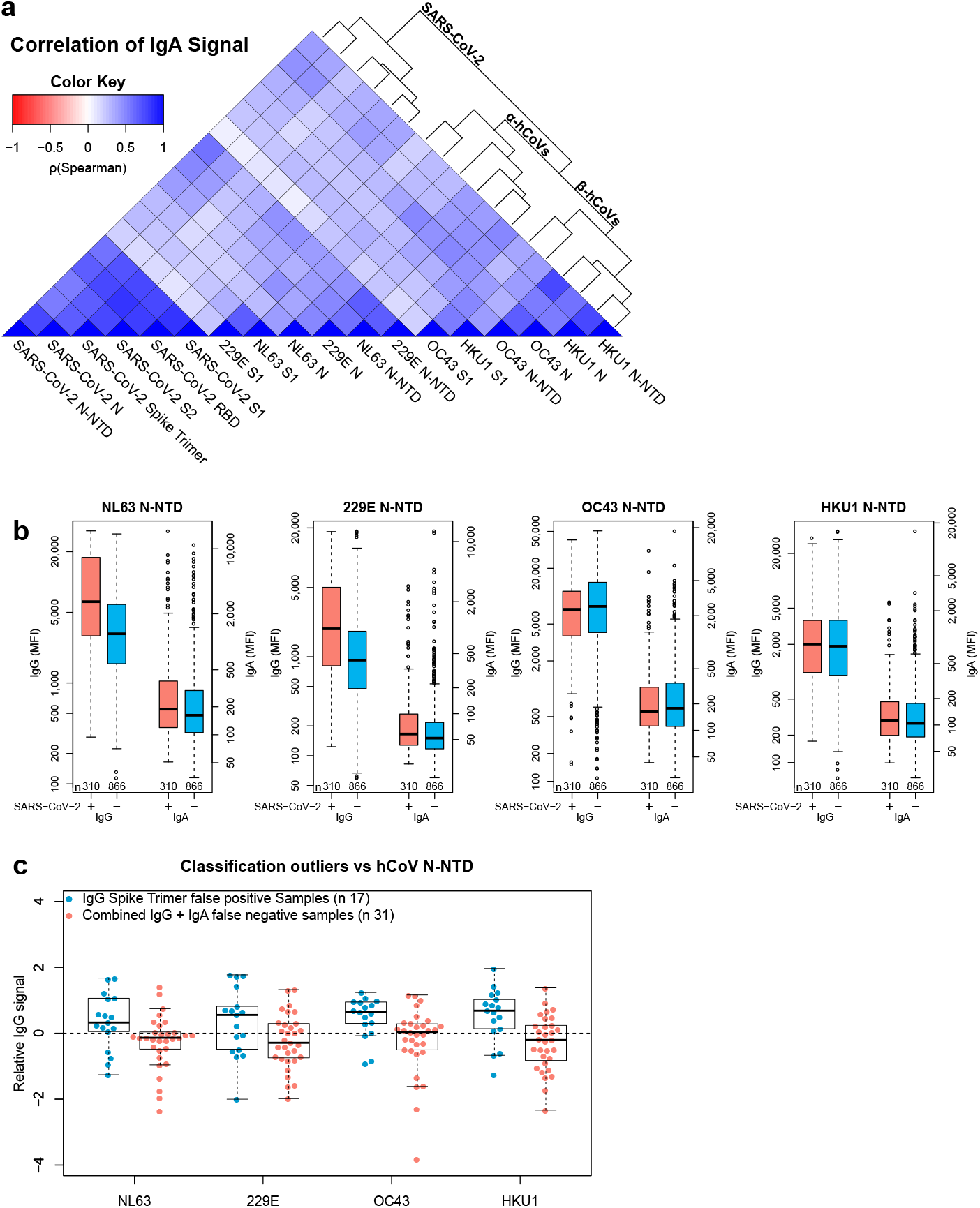
**a**, Correlation of IgA response for the entire sample set (n=1176) is visualized as heatmap based on Spearman’s ρ coefficient; dendrogram on the right side displays antigens after hierarchical clustering was performed. **b**, Immune response (IgG and IgA) towards hCoV N-NTD proteins are presented as Box-Whisker plots of sample MFI on a logarithmic scale for SARS-CoV-2-infected (red, n=310) and uninfected (blue, n=866) individuals. Outliers determined by 1.5 times IQR of log-transformed data are depicted as circles. **c**, Relative levels of IgG-specific immune response towards hCoV N-NTD proteins are presented as Box-Whisker plots / stripchart overlays of log-transformed and per-antigen scaled and centred MFI for the sample subsets of Spike Trimer false positives (blue, n=17) and combined IgG + IgA false negatives (red, n=31).

**Extended Data Table 1.** Intra-and inter-assay variance were determined by repeated measurement of QC samples and blank sample as replicates on one plate and in duplicates over 17 plates, respectively. Standard deviation relative to mean (%CV) is given for each antigen. A limit of detection (LOD) was calculated from 24 blank sample replicates on the same plate as the mean MFI + 3 times standard deviation.

|  |  |  | SARS-CoV-2 |  |  |  |  |  | hCoV NL63 |  |  | hCoV 229E |  |  | hCoV OC43 |  |  | hCoV HKU1 |  |  |
| --- | --- | --- | --- | --- | --- | --- | --- | --- | --- | --- | --- | --- | --- | --- | --- | --- | --- | --- | --- | --- |
|  |  |  | Spike Trimer | RBD | S1 | S2 | N | N-NTD | S1 | N | N-NTD | S1 | N | N-NTD | S1 | N | N-NTD | S1 | N | N-NTD |
| Inter-assay variance (%CV)<br>n = 34,<br>duplicates,<br>17 plates | IgG | QC1 | 3.7 | 3.3 | 3.4 | 3.7 | 2.8 | 7.4 | 3.5 | 3.2 | 4.4 | 3.3 | 2.8 | 5.2 | 3.1 | 6.0 | 4.4 | 3.4 | 4.7 | 5.4 |
|  |  | QC2 | 4.1 | 4.6 | 6.9 | 3.4 | 5.3 | 4.8 | 3.0 | 2.2 | 6.3 | 2.4 | 2.1 | 6.7 | 2.7 | 4.5 | 2.3 | 2.7 | 5.1 | 2.8 |
|  |  | QC3 | 3.4 | 2.4 | 2.3 | 3.6 | 2.1 | 4.6 | 3.1 | 2.5 | 3.5 | 2.9 | 2.0 | 4.7 | 2.9 | 6.4 | 4.6 | 3.2 | 3.2 | 3.5 |
|  |  | Blank | 5.4 | 5.6 | 6.7 | 6.4 | 5.6 | 6.1 | 6.3 | 7.1 | 5.7 | 9.1 | 6.1 | 6.1 | 5.6 | 7.3 | 4.1 | 4.9 | 6.1 | 8.3 |
|  | IgA | QC1 | 3.9 | 4.6 | 4.9 | 4.0 | 5.1 | 5.0 | 4.2 | 3.6 | 3.9 | 4.0 | 5.3 | 7.4 | 4.3 | 7.4 | 5.0 | 4.2 | 6.0 | 5.0 |
|  |  | QC2 | 4.6 | 5.0 | 5.1 | 3.9 | 3.9 | 4.2 | 3.7 | 2.4 | 7.6 | 2.9 | 2.2 | 6.0 | 4.1 | 16.4 | 4.2 | 4.3 | 5.5 | 3.9 |
|  |  | QC3 | 3.9 | 3.8 | 4.5 | 3.4 | 3.4 | 4.8 | 3.9 | 2.8 | 4.0 | 3.0 | 5.1 | 4.5 | 3.6 | 6.1 | 4.1 | 3.7 | 4.5 | 5.1 |
|  |  | Blank | 6.7 | 5.3 | 8.2 | 6.3 | 5.3 | 5.3 | 3.3 | 5.0 | 5.0 | 6.7 | 7.0 | 6.1 | 5.3 | 7.1 | 4.7 | 6.0 | 6.8 | 6.3 |
| Intra-assay variance (%CV)<br>n = 24 | IgG | QC1 | 2.5 | 1.9 | 2.0 | 2.1 | 1.8 | 2.1 | 2.4 | 1.7 | 2.8 | 2.0 | 2.7 | 3.2 | 1.9 | 2.0 | 2.2 | 2.7 | 2.4 | 2.2 |
|  |  | QC2 | 5.9 | 4.3 | 4.1 | 2.8 | 2.7 | 3.2 | 1.9 | 1.9 | 2.6 | 2.0 | 2.2 | 2.7 | 2.2 | 1.6 | 2.1 | 2.5 | 3.1 | 2.5 |
|  |  | QC3 | 1.6 | 4.3 | 5.1 | 1.9 | 1.9 | 4.5 | 4.2 | 1.7 | 3.2 | 3.3 | 4.1 | 5.7 | 3.2 | 3.1 | 5.5 | 5.6 | 6.0 | 8.4 |
|  |  | Blank | 6.0 | 5.6 | 5.2 | 5.8 | 5.2 | 4.2 | 5.0 | 4.8 | 4.8 | 7.3 | 6.2 | 6.3 | 6.5 | 6.2 | 4.4 | 6.1 | 6.0 | 6.2 |
|  | IgA | QC1 | 2.5 | 3.3 | 5.2 | 3.8 | 3.7 | 4.2 | 3.2 | 2.3 | 2.2 | 2.0 | 4.8 | 4.7 | 2.9 | 4.7 | 3.4 | 3.3 | 4.5 | 4.3 |
|  |  | QC2 | 4.8 | 5.7 | 5.7 | 3.2 | 4.1 | 4.3 | 3.4 | 2.0 | 5.7 | 2.1 | 2.1 | 6.1 | 3.0 | 3.1 | 1.9 | 3.9 | 6.4 | 3.8 |
|  |  | QC3 | 3.1 | 4.7 | 5.5 | 3.0 | 4.1 | 4.4 | 3.7 | 2.7 | 3.7 | 2.7 | 5.4 | 5.8 | 2.6 | 4.1 | 3.1 | 2.4 | 4.5 | 4.3 |
|  |  | Blank | 5.8 | 5.3 | 6.3 | 5.0 | 5.4 | 5.5 | 4.5 | 5.6 | 5.3 | 7.2 | 6.3 | 7.0 | 6.7 | 9.5 | 7.3 | 5.2 | 8.8 | 7.0 |
| LOD* (MFI)<br>n = 24 | IgG |  | 32 | 26 | 23 | 29 | 38 | 33 | 29 | 28 | 26 | 65 | 35 | 24 | 33 | 30 | 33 | 37 | 33 | 25 |
|  | IgA |  | 31 | 26 | 26 | 27 | 37 | 32 | 57 | 28 | 40 | 28 | 36 | 22 | 35 | 28 | 32 | 33 | 39 | 28 |

\*Blank mean MFI + 3 \* sd

**Extended Data Table 2.**
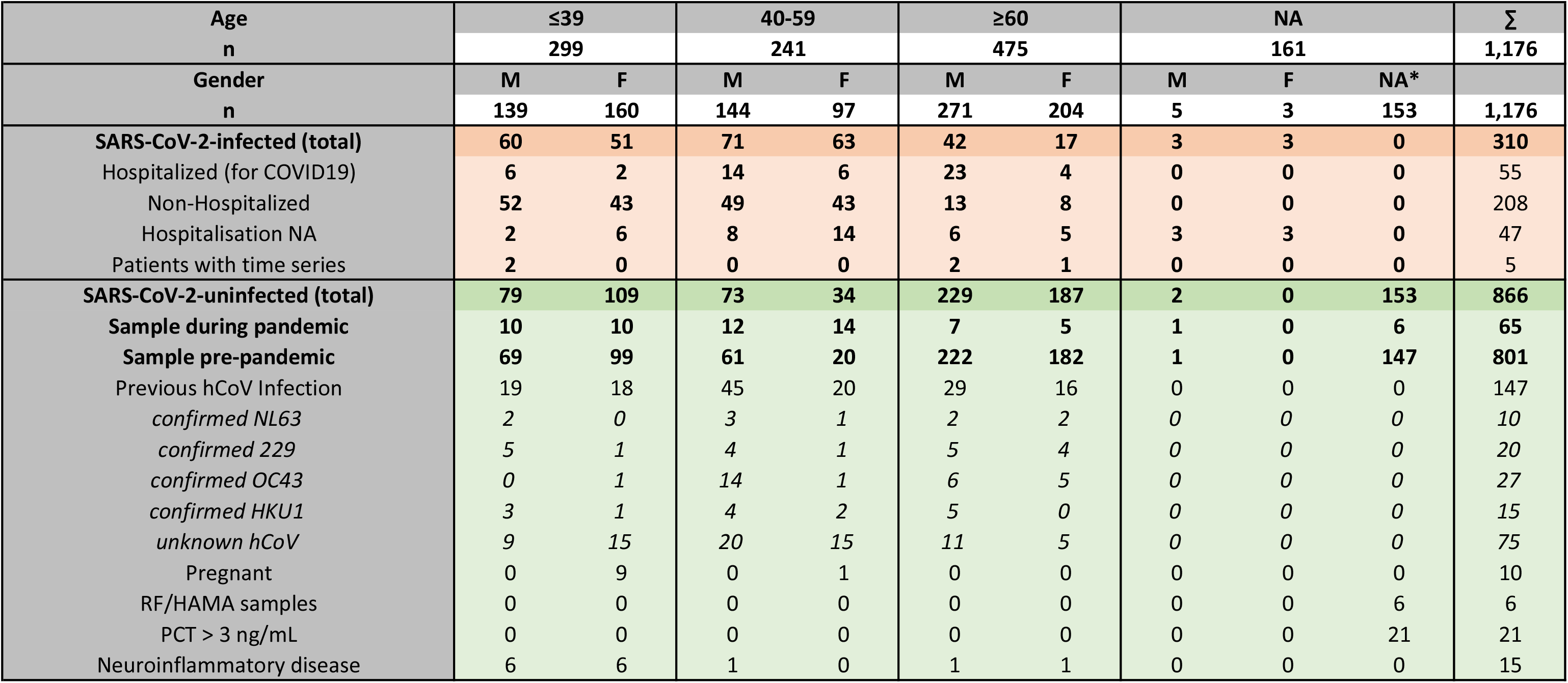
Complete overview of study sample set. Samples are divided into columns by age groups and gender. NA: Information was not available. Samples from SARS-CoV-2-infected donors are further split up by hospitalization status. Age and gender of patients from which multiple samples were available for time course analyses are indicated. SARS-CoV-2-uninfected samples are further divided into samples drawn during the pandemic, which was defined as all samples taken on 01.01.2020 or later, and pre-pandemic samples. 147 samples with previous hCoV infection were included in the SARS-CoV-2-uninfected group. Detailed diagnosis of hCoV subspecies is indicated where available. Other sample conditions for special groups of uninfected samples are listed.

